# Transcriptomic Profiling on Localized Gastric Cancer Identified *CPLX1* as a Gene Promoting Malignant Phenotype of Gastric Cancer and a Predictor of Recurrence after Surgery and Subsequent Chemotherapy

**DOI:** 10.1101/2021.03.25.21254204

**Authors:** Haruyoshi Tanaka, Mitsuro Kanda, Dai Shimizu, Chie Tanaka, Norifumi Hattori, Yoshikuni Inokawa, Masamichi Hayashi, Goro Nakayama, Yasuhiro Kodera

## Abstract

Localized gastric cancer (GC) becomes fatal once recurring. We still have room for improving their prognoses. Firstly, a transcriptomic analysis was done on surgically resected specimens of 16 patients with UICC stage III GC who underwent curative gastrectomy and adjuvant oral fluoropyrimidine monotherapy. Four of them were free from disease for longer than 5 years, and the others experienced 15 metachronous metastasis at either liver, peritoneum, or distant lymph nodes (n = 4 each) within 2 years after surgery. *CPLX1* was identified as a novel oncogene candidate for recurrence among 57,749 genes. Secondary, we tested alteration of malignant phenotypes including drug resistance of gastric cancer cell lines by small interfering RNA-mediated *CPLX1* knockdown. Inhibiting *CPLX1* expression decreased the proliferation, motility, and invasiveness of GC cells, and increased apoptosis and sensitivity to fluorouracil. Thirdly, we validated the clinical significance of *CPLX1* expression in GC by quantitative RT-PCR on 180 primary gastric cancer tissues of which patients underwent gastric resection for stage II and III GC without preoperative treatment between 2001 and 2014. Increased expression of *CPLX1* mRNA in gastric cancer tissues correlated with worse prognoses and was an independent risk factor for peritoneal recurrence in subgroups receiving adjuvant chemotherapy. *CPLX1* may represent a biomarker for recurrence of gastric cancer and a target for therapy.

**Brief description:** Transcriptomic analysis identified CPLX1 gene as a novel oncogene candidate for gastric cancer. CPLX1 may promote epithelial-mesenchymal transition and evading apoptosis of gastric cancer cells even under a cytotoxic agent, and also be a predictor for recurrence after surgery for UICC Stage II-III gastric cancer.

## Introduction

Gastric cancer represents the third most leading cause of death among malignancies.^1,2^ Although the 5-year overall survival rates vary from 20% to 50% depending on region, worldwide trends improved little or rather flat in this decade.^1^ At early stages, most cases of gastric cancer can be cured by surgical eradication.^3^ In order to improve outcomes of advanced, yet localized gastric cancer, it is critical how to minimize the risk of metachronous metastases.^4^ Perioperative chemotherapy or chemoradiotherapy were widely performed as standard therapy for these locally advanced diseases.^5^ However, there is no international consensus on regimens, and they often fail to prevent cancer from recurring.^4,6^

It is considered that recurring cancers have had already malignant phenotypes in primary lesion, such as invasiveness in peritoneal cavity or circulation, or cell survival under anoikis, or resistance to cytotoxic agents. Cancer cells recurring theoretically should have been separated from primary cancer tumors at the time of surgery. Once cancer cells detach from the primary lesion and are disseminated into the blood vessel, lymph vessel, or peritoneal cavity, they are simultaneously exposed to various selective pressure, such as hypoxia and anoikis.^7^ They would even be exposed to the induction of cytotoxic agents before long, and most disseminated cancer cells are considered to be extinct. Only subclones having certain malignant phenotypes could survive under selective pressure until their outgrowth.

Global genomic, epigenomic, and transcriptomic profiling, machine learning, and high-throughput biological screening with a bioinformatic approach have paved the way towards new insights on molecules demonstrating oncogenic phenotype including multidrug resistance.^8^ In particular, global profiling approaches possess the potential for us to discover novel therapeutic targets and biomarkers.^9,10^ However, many of these profiling seems to compare in terms of the UICC stages at the time of sampling, instead of respect to their therapeutic strategies and their outcomes of recurrent status. We hypothesized that primary cancer that recurs after surgery and adjuvant chemotherapy must have a divergent gene profile from those cases without recurrence. Here, we conducted next-generation RNA sequencing (RNA-seq) on gastric cancer limited to UICC-Stage III undergoing adjuvant chemotherapy with an oral fluorinated pyrimidine to identify the *CPLX1* as a driver oncogene candidate. *CPLX1* is a member of the Complexin/Synaphin family playing a key role in synaptic vesicle exocytosis, and dysfunction of the *CPLX1* is associated with Developmental and epileptic encephalopathy-63.^11^ To the best of our knowledge, however, this is the first study to have revealed malignant roles of *CPLX1* in the outcome of gastric cancer.

## Materials and methods

### Sample collection

For transcriptomic analysis, surgically resected specimens of 16 patients with stage III gastric cancer who underwent curative gastrectomy and adjuvant S-1 monotherapy were selected. These patients were categorized into two groups: those who were free from disease for longer than 5 years (n = 4) versus those who suffer from the liver, peritoneal, or distant nodal recurrences (n = 4 each) within 2 years after surgery. Global expressional data of the external dataset was obtained from KMplotter (https://kmplot.com/analysis/).^12^ Particularly, the data were retrieved from the GSE62254, GSE15459, GSE22377, GSE62254, GSE29272, GSE38749 limited to UICC stage II-III.

Gastric cancer cell lines (AGS, RRID is CVCL_0139; GCIY, CVCL_1228; IM95, CVCL_2961; KATOIII, CVCL_0371; MKN1, CVCL_1415; MKN7, CVCL_1417; MKN45, CVCL_0434; MKN74, CVCL_2791; N87, CVCL_1603; NUGC2, CVCL_1611; NUGC3, CVCL_1612; NUGC4, CVCL_3082; OCUM1, CVCL_3084; and SC-6-JCK, CVCL_F953) and the non-tumorigenic tubular epithelial cell line FHs74 (CVCL_2899) were obtained from the American Type Culture Collection (Manassas, VA, USA), the Japanese Collection of Research Bio Resources Cell Bank (Osaka, Japan), or were established at our institute. All cell lines were tested for mycoplasma, analyzed using the short tandem-repeat-polymerase chain reaction (PCR) method, and authenticated by the JCRB Cell Bank on June 30th, 2015. Cell lines were cultured at 37°C in RPMI 1640 medium (Sigma Aldrich, St. Louis, MO, USA) supplemented with 10% fetal bovine serum in an atmosphere containing 5% CO_2_.

Primary gastric cancer tissues were collected from 180 patients who underwent gastric resection of stage II and III gastric cancer without preoperative treatment at Nagoya University Hospital between 2001 and 2014. Since 2006, adjuvant oral fluoropyrimidine monotherapy has been recommended to all patients with stage II and III gastric cancer unless contraindicated. Primary gastric cancer tissue specimens were classified histologically according to the 7th edition of the Union for International Cancer Control (UICC) classification system and Japanese classification of gastric carcinoma (3rd edition) by two pathologists. This study conformed to the ethical guidelines of the World Medical Association Declaration of Helsinki Ethical Principles for Medical Research Involving Human Subjects. This study was approved by institutional review board at Nagoya University, Japan. Written informed consent was obtained from all patients for the use of clinical samples and data.

### Global expression profiling analysis

Library preparation for RNA-seq was performed as described previously.^13^ To be brief, total RNAs (10 μg) isolated from each of the surgically resected gastric tissues by the RNeasy Mini Kit (Qiagen, Hilden, 15 Germany). Only samples that passed the quality test by Agilent 2100 Bioanalyzer (Agilent, Santa Clara, CA, USA) were analyzed. Poly (A)-RNA was purified using oligo-dT beads, from which the cDNAs were synthesized, followed by adding 3’A. On the cDNAs, adaptors were ligated to be amplified by cycles of PCR. Detailed RNA sequencing protocol was also described previously.^13^ Briefly, two runs per sample were performed with the paired-end method on the HiSeq sequencing system (Illumina, San Diego, CA, USA). Then read mapping and quantification of gene expression were done. Mapping of the trimmed read was performed applying TopHat with annotations of GRCh37. Normalization of read counts of each gene was done with the TMM method, and the significance of their differences between the two groups (free from cancer for more than 5 years vs. early recurring cancer) were calculated with the Exact test in edgeR and TCC packages in R. In these packages, the moderated tagwise dispersion of the normalized read counts estimated by the quantile-adjusted conditional maximum likelihood method were used.^14^ *P* values were adjusted by applying Benjamini-Hochberg method with a false discovery rate of 0.5.

### Quantification of CPLX1 expression by real-time reverse transcriptional PCR (qRT-PCR)

*CPLX1* mRNA expression levels were determined using qRT-PCR as described previously.^15^ Briefly, total RNAs (10 μg) were isolated with the RNeasy Mini Kit (Qiagen) from each of the gastric cancer cell lines and surgically resected gastric cancer tissues, and their qualities were checked by measuring optical density (OD). cDNAs were generated from the RNAs and were amplified using specific PCR primers for *CPLX1* (Supplemental Table S1). A Sybr-Green PCR core reagents kit (Applied Biosystems, Foster City, CA, USA) was used to perform qRT-PCR. Initial denaturation at 95°C for 10 min, 50 cycles 10 at 95 °C for 10 s, and 60 °C for 30 s. All clinical samples were tested in technical triplicate.

### Knockdown of CPLX1 expression using small interfering RNA (siRNA)

*CPLX1*-specific siRNA sequences were designed using two web-based tools; iScore Designer and siDirect; https://www.med.nagoya-u.ac.jp/neurogenetics/i_Score/i_score.html and http://sidirect2.rnai.jp/. siRNA specific for *CPLX1* or a control non-specific siRNA (40 nM of each) (Bioneer, Daejeon, Korea) were transiently transfected into gastric cancer cells using Neon® transfection system for OCUM1 or LipoTrust EX Oligo (Hokkaido System Science, Sapporo, Japan) for others. Seeding for all assays was done 24 h after transfection, which was performed on the next day of passage.

### Cell proliferation and apoptosis assay

Cell proliferation was evaluated by measuring sequential cell viability using the Premix WST-8 Cell Proliferation assay kit (Dojindo Inc., Kumamoto, Japan) as described previously.^16^ Cancer cell lines (5×10^3^ cells/well each) were seeded into 96-well plates. The OD of the solution at 450 nm in each well was measured 2 hours after the addition of 10 μL of WST-8, and the fold-change in OD from the day of seeding (day 0) was evaluated. The assay was done in a technical quintuplicate.

The detection of apoptotic status was evaluated with annexin-V staining as described previously.^17^ To be brief, cells are diluted in annexin-binding buffer and 10 µL of annexin-V conjugate (Thermo Fisher Scientific). The cells were mounted on slides, and the apoptotic cells labeled fluorescently were counted under ×200 magnification on BZ9000 (Keyence, Osaka, Japan). UV-irradiated cells for 8h followed by 16h incubation in the normal condition was used as positive control.

### Cell migration and invasion assay

The migration of AGS cells was tested in technical duplicate as described as previously, using Culture-Insert 2 Well in µ-Dish 35 mm (ibidi GmbH, Martinsried, Germany).^18^ Briefly, after 16 h passed from the seeding of AGS cells, the inserts were removed to establish cell-free gaps. The distances of cells were measured from both edges at 20 points per well. The mean widths of the cell-gap at the time of zero were adjusted to 580µm for equalization.

The ability of AGS cells to invade Matrigel matrix was determined in technical duplicate as described previously, using BioCoat Matrigel invasion chambers (BD Biosciences, Bedford, MA, USA).^19^ AGS cells (5×10^4^ cells/well) were seeded into the upper well of the chamber in serum-free RPMI 1640 medium. After 48 h, cells on the lower surface of the membrane were fixed, stained with Giemsa. The gross views of cell invasion were evaluated under ×40 magnification.

### Drug sensitivity assay

To test the influence of *CPLX1* expression levels and knockdown effect on the resistance of gastric cancer cells to fluorouracil, we conducted drug sensitivity test as described previously.^20^ To be brief, AGS, KATOIII, MKN1, MKN45, N87, NUGC4, OCUM1 cells (5×10^3 per well, 5 wells for each condition) were treated for 96 or 120 h with fluorouracil at final concentrations of 0 to 256mg/L. Cell viabilities were measured using the Cell Counting Kit-8 (Dojindo). In regard to data analysis, we evaluated drug sensitivity with normalized growth rate inhibition (GR) metrics, to minimize the perturbation of growth rate by treatment of siRNA.^21^GR value at a time (*t*) under a drug at a concentration (*c*): GR (*c,t*) = 2^k(*c,t*)/k(0)^ – 1, where k(*c,t*) is the growth rate of drug-treated cell and k(0) is that of the untreated control cells. That means that the GR value is the ratio of treated cell’s growth rate to untreated cell’s growth rate, both of which normalized by single-cell division. GR = 0 means complete cytostasis and GR_50_ is the drug concentration where GR = 0.5.

### Exploring cancer-related genes cooperatively expressed with CPLX1 and pathway analysis

To explore genes that potentially interact with *CPLX1*, we employed two array-based gene profiling assays. To see coexpression with *CPLX1* mRNA expressional levels of EMT-related genes of 14 gastric cancer cell lines were quantified with the Human Epithelial to Mesenchymal Transition RT^2 Profiler PCR Array (Qiagen) following manufacture’s protocol.^22^ In addition, to explore which pathways *CPLX1* is involved in, the PathScan Intracellular Signaling Array Kit (Product No. 7323, Chemiluminescent Readout) (Cell Signaling Technology, Danvers, MA, USA), which is based on the sandwich immunoassay principle, was applied following manufactures protocol.^23^ KATOIII cells without treatment, treated by negative control siRNA (siControl) and si*CPLX1* were harvested to be tested. Antigen-Antibody complexes were detected with HRP-linked streptavidin through ImageQuant LAS 4000 (GE Healthcare, Chicago, IL, USA).

### Statistical analysis

The significance of the difference between variables of two groups was assessed using the Mann-Whitney test or the Student’s t-test for two valuables depending on their distribution. The Kruskal-Wallis test for three or more clinical valuables and pairwise t-test were applied for experimental values. Fisher’s exact test was used to analyze categorical data of two groups. Overall survival and disease-free survival rates were calculated using the Kaplan-Meier method, and the differences between survival rates were evaluated by the Wilcoxon test. A *P* value <0.05 was considered statistically significant. To calculate GR values, the GRmetrics package was applied.^21^ All statistical analyses except for some pipelines of the RNA-seq as mentioned above were performed with R software (The R Foundation for Statistical Computing, Vienna, Austria) version 4.0.1.

## Results

### RNA sequencing identified CPLX1 as a study subject

Global expression profiling was performed to compare the expression levels of 57,749 genes of two groups; recurrent at liver, peritoneum, or distant lymph node vs. without recurrent over 5 years, where one sample of the case that recurred at the liver was excluded due to low quality. The total RNA had adequate quality as follows; 21.8 million pairs of mean reads were yielded per sample and the mean rate of ≥Q30 was 95.0%. We tested 57,749 genes if they were differentially expressed between postoperative cases with and without recurrence (Fig. 1a). Among 305 differentially expressed genes, *CPLX1* was one of 12 genes of protein-coding and upregulated in the early recurrent group (Fig. 1a-c). As the predictive performance of *CPLX1* was affirmed with external data sets (Supplemental Fig. 2a, 2b), we chose *CPLX1* as a candidate for oncogene and to be subject to further investigation.

**Fig. 1.**
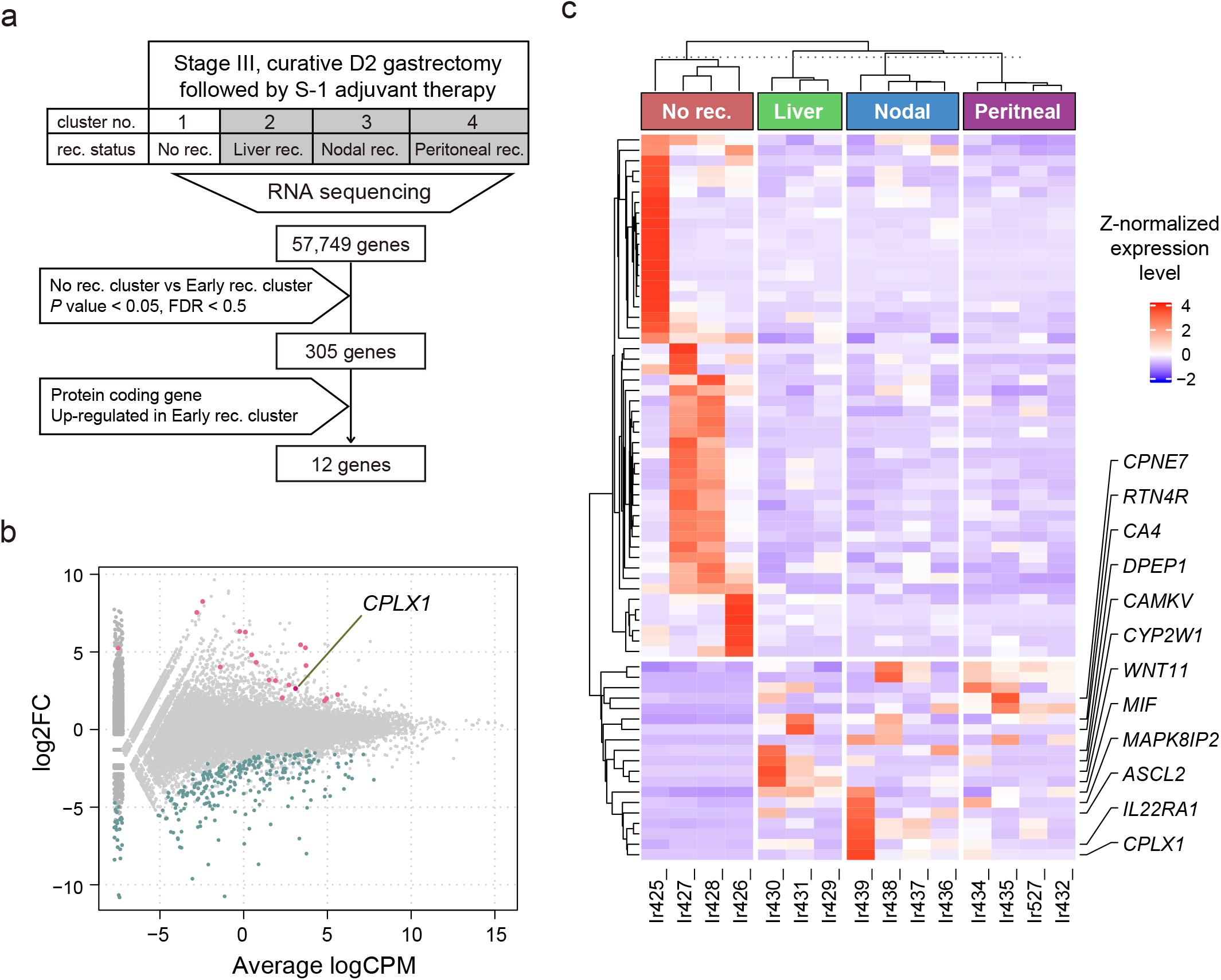
CPLX1 was identified as a putative oncogene to promote the recurrence of gastric cancer. (a) Extraction diagram demonstrating candidate genes identified by transcriptome analysis. Expressions of 57,749 genes were compared between those who were free from recurrence and those who experienced recurrences of gastric cancer within 2 years. (b) Log-ratio and mean average (MA) plot of expression levels between no recurrence group and early recurrence group yielded twelve candidates of oncogenes. Differentially expressed genes (DEGs) were indicated by magenta (up-regulated in the early recurrence group) and green (down-regulated). (c) RNA expressional heatmap of surgically resected specimens of stage III gastric cancer and adjuvant oral fluoropyrimidine monotherapy. Up-regulated, protein-coding 12 DEGs were listed on right. FC, fold change; CPM, counts per million.

### Inhibition of CPLX1 decreases malignant phenotypes of gastric cancer cells

Expressional levels of *CPLX1* mRNA were heterogeneous among gastric cancer cell lines and not detected in FHs74 (Fig. 2a). Four siRNAs were designed to determine the effect of knockdown on phenotypes (Supplemental Table S1). The knockdown effective ratio of si*CPLX1*-treated cells compared to untransfected cells ranged 0.168 – 0.266, 0.341 – 0.418, and 0.278 – 0.342, for OCUM1, N87, and KATOIII cells respectively (Fig. 2b). The proliferation of AGS, MKN1, OCUM1, and KATOIII cells treated with si*CPLX1* was significantly reduced compared to the untreated cells and those treated by siControl (Fig. 2c). At gross appearance through a phase-contrast microscope, we observed that AGS cells transfected by si*CPLX1* had cytoplasmic vacuolization in their cell cytoplasm (Fig. 2d). That seemed to indicate they were toward cell death^24^ so that we sought to quantify *CPLX1* promote apoptotic evasion. The proportions of annexin-V positive apoptotic cells were significantly increased by inhibition of *CPLX1* expression on AGS, KATOIII, MKN1, and N87 cells (Fig. 2e, Supplemental Fig. 2a). Additionally, to observe if si*CPLX1* hampers cancer cells from epithelial-mesenchymal transition (EMT), we performed migration assay and invasion assay on AGS cells. Migrating distances of AGS cells were significantly reduced by si*CPLX1* (Fig. 2f). Also, fewer AGS cells treated by si*CPLX1* passed through Matrigel chambers compared to AGS untreated or treated by siControl (Fig. 2g)

**Fig. 2.**
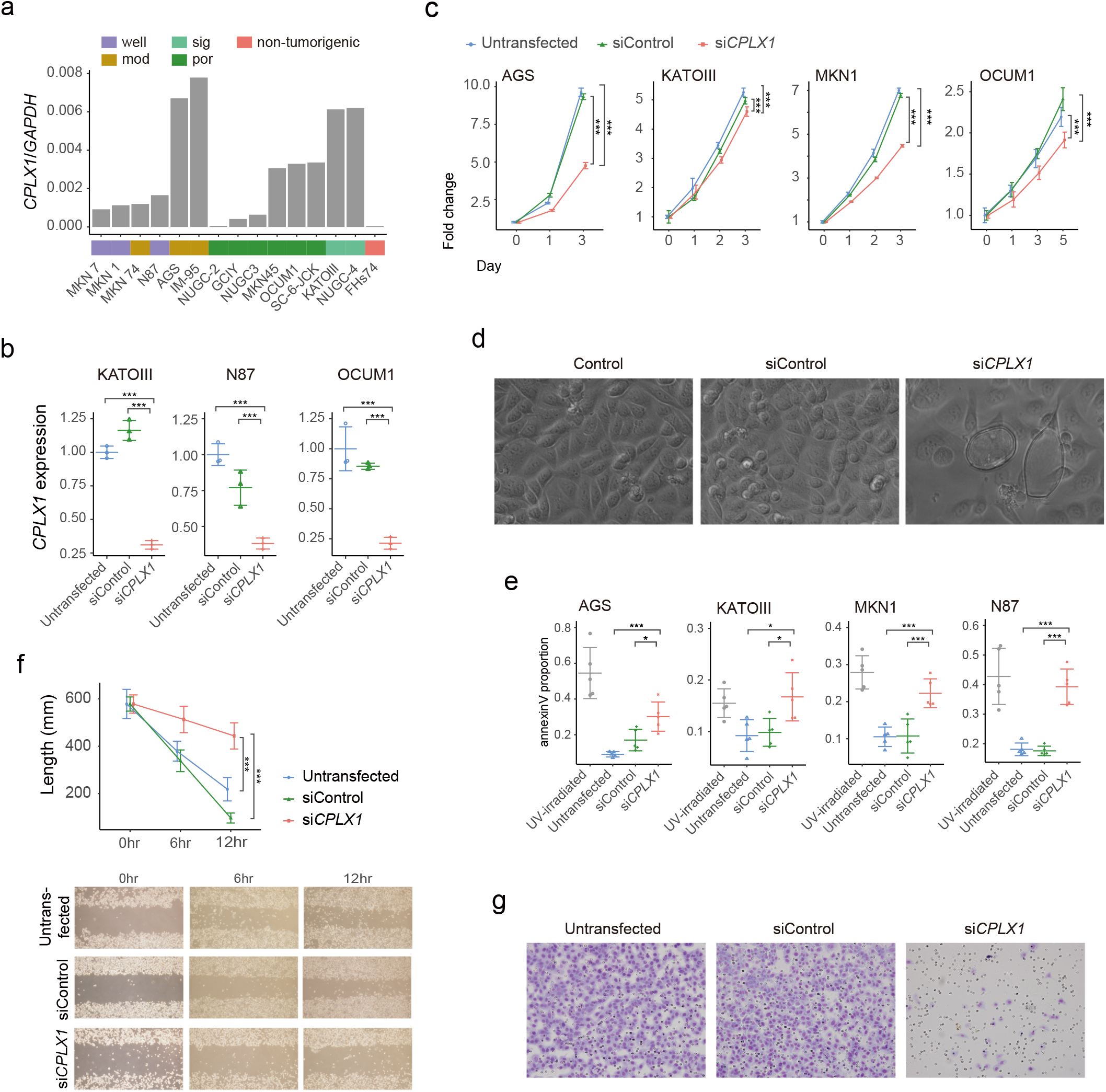
CPLX1 promotes malignant phenotypes of gastric cancer cell lines. (a) CPLX1 expression levels of mRNA on 14 gastric cancer (GC) cell lines and FHs74, nontumorigenic tubular epithelial cell line. (b) CPLX1-knockdown effect on three GC cell lines at mRNA levels. (c) Proliferation assay comparing those four GC cell lines between untreated, transfected by siControl and siCPLX1. Apoptotic cell observation (d) at cultured on dishes through a phase-contrast microscope, and (e) by staining with annexin-V. Those four GC cell lines were compared between untreated, treated by siControl, treated by siCPLX1. Cells treated by UV-irradiation were positive controls. (f) Migration assay and (g) invasion assay on AGS cells. *P < 0.05, **P < 0.01, ***P < 0.001.

To test the possible role of *CPLX1* in chemotherapy-resistant, sensitivities to fluorouracil were tested on seven cell lines (Supplemental Fig. S2b). Although insignificant, there were weak correlations between the area under the curves of dose-response and *CPLX1* expression levels of these 7 cell lines (Supplemental Fig. S2c). *CPLX1* inhibition increased sensitivities to fluorouracil of KATOIII, OCUM1, MKN1, and AGS cells (Fig. 3a). Regarding AGS cells, statistical modeling by GR metrics failed to generate a recognizable curve for AGS cells treated by si*CPLX1*, so that conventional dose-response curves and half-maximal inhibitory concentration (IC_50_) values were presented.

**Fig. 3.**
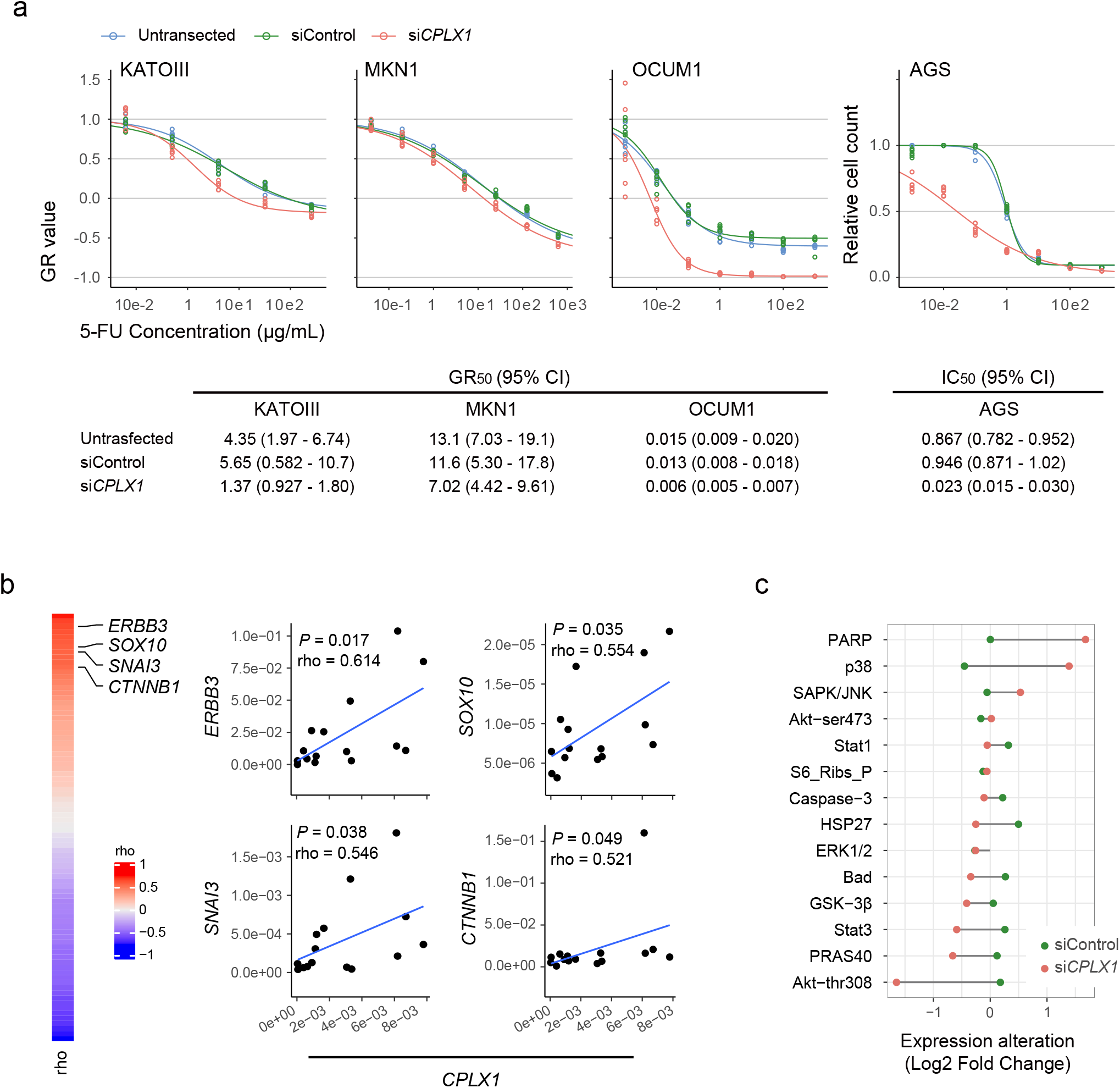
Effect of *CPLX1 i*nhibition on drug sensitivity and exploring co-expressed genes with CPLX1. (a) Drug sensitivity test comparing between untransfected, and transfected by siContorl and transfected by siCPLX1 on four different cells. GR values indicate values calculated by normalized growth rate inhibition (GR) metrics. (b) PCR-based EMT-related 84 genes profiling to test co-expression with CPLX1 among 14 GC cell lines. Significances of coefficients were tested with Spearman correlation test and coefficients degree were represented by rho (ρ) values. (c) Pathway analysis with PathScan, sandwich ELISA based immunoassay comparing KATOIII cells untreated, treated by siControl, and siCPLX1. All values were presented with Log2 fold change (Log2FC) from untreated KATOIII cells. Only targets of which absolute values of log2FCs (i.e. |log2FC|) between untransfected and transfected by siControl are less than 0.5 are presented.

We further explored genes that potentially interact with *CPLX1*. Array-based PCR assay revealed that *ERBB3, SOX10, SNAI3*, and *CTNNB1* were cooperatively expressed at a moderate level, of which rho ranged between 0.521 to 0.636, with *CPLX1* among 14 gastric cancer cell lines (Fig. 3b). Also, array-based protein phosphorylation analysis showed si*CPLX1* inhibited phosphorylation of Akt at thr308 and PRAS40 at Thr246 were weakly inhibited compared to both untransfected and transfected with siControl in KATOIII cells. Conversely, upregulation of phosphorated p38 at Thr180/Tyr182 and cleaved PARP at Asp214 were observed. (Fig. 3c)

### Overexpression of CPLX1 in tumor tissues predicts recurrence of gastric cancer

Forty-five cases of 180 stage II-III gastric cancer patients were classified into a high *CPLX1* expression group according to the upper quantile value of the whole cohort (*CPLX1*/*GAPDH* > 0.06882). *CPLX1* expression levels were independent of all clinicopathological variables of 180 patients of stage II-III gastric cancer except for tumor diameter and main tumor locations (Table 1). High *CPLX1* expression group had a poor overall-survival: 5-year survival rates were as low as 0.593 (95% confidential interval: 0.460 – 0.763), compared to 0.744 (0.665 – 0.832) for low *CPLX1* group (*P* = 0.025, Fig. 4a). High *CPLX1* expression group also had shorter recurrence-free survival; 3-year survival rates were 0.514 (0.385 – 0.687), compared to 0.702 (0.628 – 0.785) for low *CPLX1* group (*P* = 0.032, Fig. 4b). To further investigate the association between *CPLX1* and resistance to adjuvant chemotherapy in terms of recurrence organs, we performed a subgroup analysis on a cohort that underwent adjuvant chemotherapy.

**Table 1.**
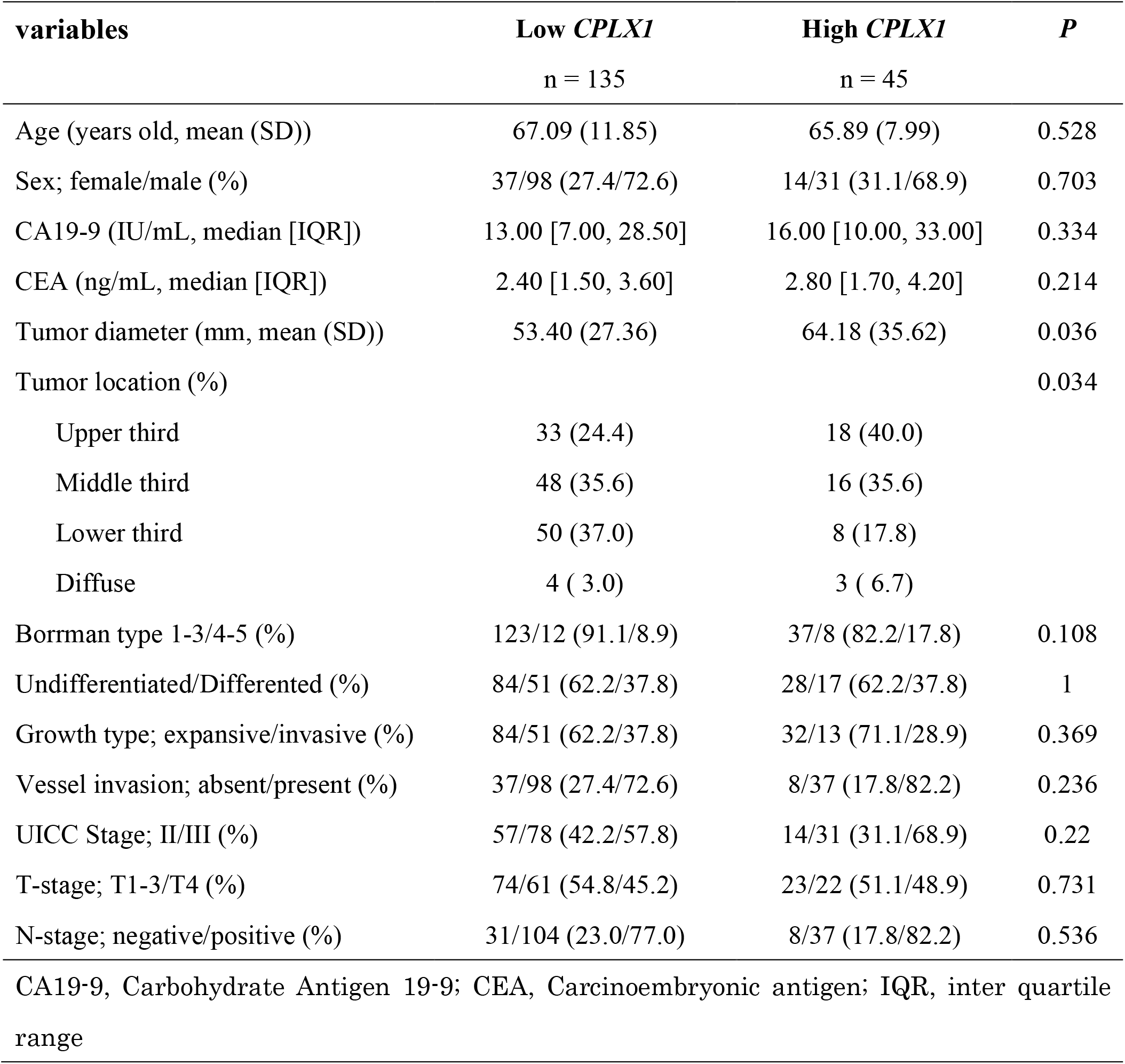
clinicopathological characteristics of stage II-III gastric cancer (n = 180)

**Fig. 4.**
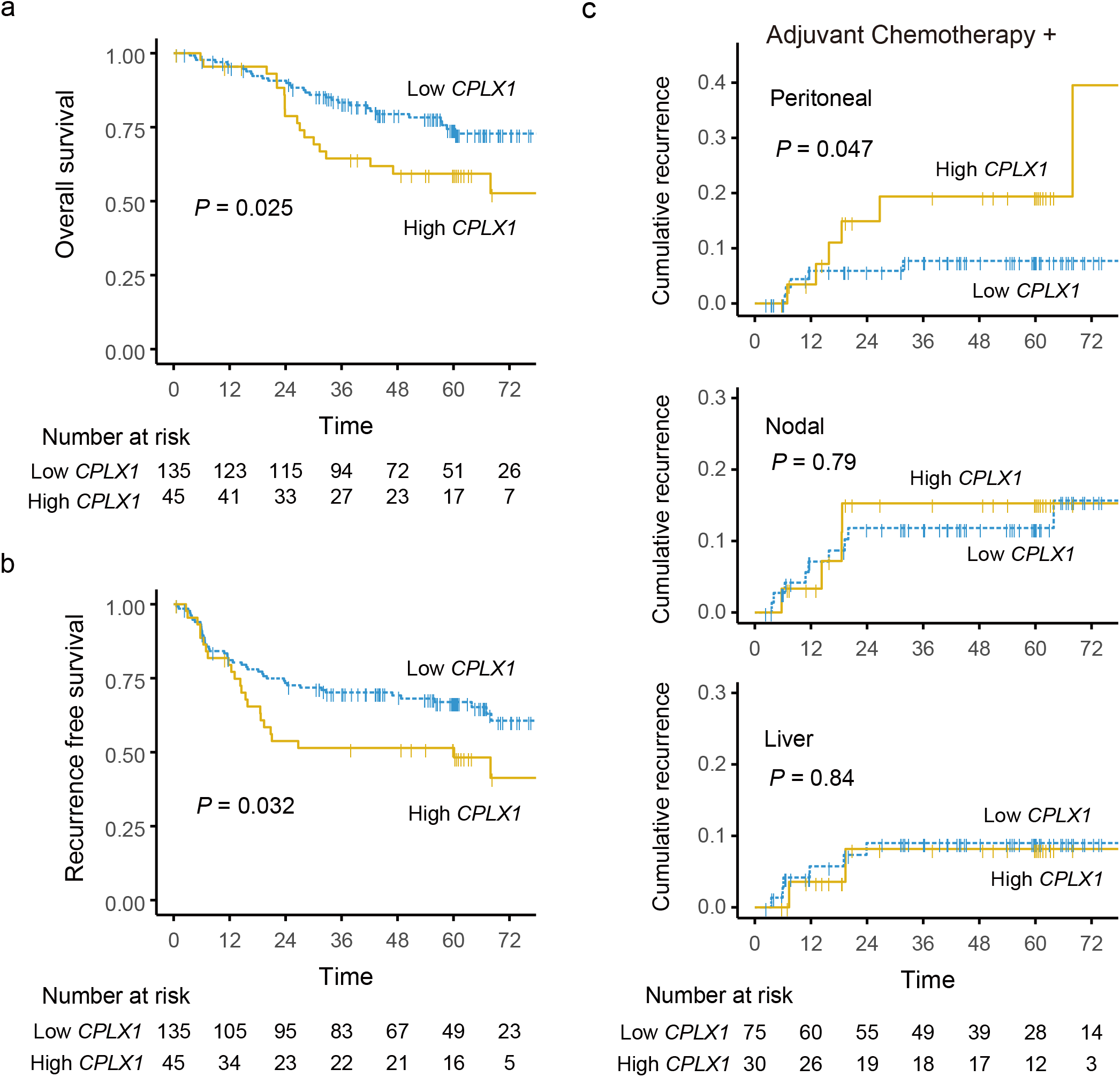
High CPLX1 expression correlates with the poor survival of patients with gastric cancer. Kaplan-Meier plots of (a) overall survival and (b) recurrence-free survival for the whole cohort (n = 180) by stratifying with CPLX1 mRNA expression (CPLX1/GAPDH). (c) Organ-specific cumulative recurrent plot subgroup receiving adjuvant chemotherapy (n = 105).

The high *CPLX1* group was likely to have a higher cumulative peritoneal recurrence rate than low *CPLX1* group (*P* = 0.047), but this is not the case for liver or nodal recurrence (Fig. 4c). The univariate Cox hazard model showed that high *CPLX1* expression was not significant but tend to be a high-risk factor for peritoneal recurrence in cohorts having chemotherapy (hazard ratio, 3.139 (0.95 – 10.33); *P* = 0.062). Since the factor tumor diameter < 50mm being a possible confounder of *CPLX1, CPLX1* was included in multivariate analysis with tumor diameter to demonstrate that high *CPLX1* expression was one of the independent risk factors for peritoneal recurrence (hazard ratio, 3.356 (1.006 – 11.2); *P* = 0.049) (Table 2).

**Table 2.** COX hazard model analysis for peritoneal recurrence after adjuvant chemotherapy

| Variable | coefficient | *P* | coefficient | *P* |
| --- | --- | --- | --- | --- |
| Age >65 years old | 0.654 (0.19 – 2.24) | 0.492 |  |  |
| Male sex | 0.622 (0.19 – 2.06) | 0.442 |  |  |
| Tumor size >50 mm | 8.403 (1.08 – 65.71) | 0.008* | 5.465 (0.69 – 43.54) | 0.109 |
| Tumor location (upper third) | 1.32 (0.35 – 4.98) | 0.688 |  |  |
| CA19-9 >37 IU/mL | 0.496 (0.063 – 3.874) | 0.461 |  |  |
| CEA >5 ng/mL | 1.155 (0.249 – 5.348) | 0.856 |  |  |
| Differentiated type | 0.422 (0.091 – 1.959) | 0.232 |  |  |
| Invasive growth type | 7.442 (1.60 – 34.54) | 0.003* | 7.531 (1.60 – 35.47) | 0.011* |
| Vessel involvement | 1.295 (0.280 – 6.00) | 0.734 |  |  |
| Tumor depth T4 (vs T1 – 3) | 2.371 (0.630 – 8.94) | 0.179 |  |  |
| Lymph node metastasis | NA (0 – Inf) | NA |  |  |
| UICC Stage III | NA (0 – Inf) | NA |  |  |
| High *CPLX1* expression | 3.139 (0.954 – 10.3) | 0.062 | 3.356 (1.01 – 11.2) | 0.049* |

## Discussion

The prognosis of gastric cancer remains unsatisfactory, and the diagnostic and therapeutic strategy for recurrence has not been developed substantially. Here, we uncovered that *CPLX1* was distinctly overexpressed in primary tumor tissues of stage III gastric cancer that recurred despite treatment with curative resection and subsequent chemotherapy with an oral fluorinated pyrimidine. Subsequent qRT-PCR analysis of 180 gastric cancer tissues validated the discovery by showing that higher *CPLX1* expression correlates with a worse prognosis especially in terms of peritoneal recurrence after adjuvant chemotherapy with oral fluoropyrimidine. Our knockdown assays suggested that *CPLX1* expression in a gastric cancer cell line promotes malignant phenotypes, such as evading apoptosis, migration, and invasion into extracellular matrix. The drug sensitivity test showed that *CPLX1* is not only a candidate biomarker predicting resistance to chemotherapy, but also may have functional aspects of resistance to fluorinated pyrimidines.

Present transcriptomic analysis was attentively designed with limit of UICC stage III gastric cancer patients who underwent adjuvant chemotherapy with fluorinated pyrimidine and whose prognosis was well-surveyed (i.e., more than 5 years for controls). This approach enabled us to identify genes associated with gastric cancer recurrence under specific circumstances. Some of 12 candidate up-regulate genes were reported as oncogenes in the literature. In particular, *ASCL2* was already reported as a promoter of chemo-resistance.^25^ *CPLX1* was chosen for further investigation as a novel oncogene candidates which has never been reported before. In general, cancer cells can obtain two different drug resistance mechanisms: primary and secondary acquired mechanisms.^8^ In the present study, we focused on the former and hypothesized that disseminated and anoikic cancer cells have a similar gene profile with the primary tumor, which would determine if they are able to survive under exposure to a cytotoxic agent.

In the present study, we applied GR metrics developed by Hafner et al. to evaluate the alteration of drug sensitivity by si*CPLX1*, instead of conventional IC value. That is because we sought to see the effect of *CPLX1* on drug sensitivity by adjusting confounders of proliferation. IC_50_ can vary drastically if control cells untreated by cytotoxic agents have different division rates among perturbations (i.e., untransfected, siControl, and si*CPLX1*) during an assay.^21^ The GR value is the ratio of treated cell growth rate to that of untreated cells which are normalized to the cell division. GR_50_ yielded by GR metrics is robust and largely independent of cell division rate and assay duration.

*CPLX1* gene resides on human chromosome 4p16.3 and codes complexin-1 and is a key protein to conduct neurotransmission via synaptic vesicle cycle (VC).^26,27^ *CPLX1* is dominantly expressed at cytosol of the central nervous system^28^. *CPLX1* clamps two plasma membranes and activates soluble N-ethylmaleimide sensitive factor attachment protein receptor (SNARE) complex, followed by being released from the unit by synaptotagmin, which allows endocytosis and exocytosis to be completed.^26,27^The dynamics and physiological roles of VC are so diverse depending on cell types that VC can promote cell migration through turnover of adhesion receptors, such as integrins, and switch points of attachment to extracellular matrix.^29-31^ It has also been reported that CPLX2, an important paralog of *CPLX1*, contributes to translocation of GLUT4 to cell membrane through VC.^32^

However, any oncogenic phenotypes or pathways mediated by *CPLX1* have not ever been reported. In the present study, several functional assays revealed that *CPLX1* promotes cell motility, cell survival, and cell proliferation even under fluoropyrimidine. These results and the literature on VC support that mechanisms in which *CPLX1* plays might be the extension of the cell border through VC. Bringing adhesion receptors onto the plasma membrane through VC may also enhance anti-anoikic and growth signal transduction.^33^ To explore molecular interactions with *CPLX1*, we performed array-based gene expressional and phosphorylation profiling. *ERBB3* is a transmembrane protein, which can initiate signals for cancer progression.^34^ *CTNNB1, SNAI3*, and *SOX10* are transcriptional factors to promote EMT.^35-37^ Through this co-expression, *CPLX1* may promote bringing a receptor to the cell surface to initiate transduction of cell growth and EMT signals. Phosphorylated PRAS40-thr246 mediates PIP3-dependent mTORC1 activation with Akt-thr308, which was modestly evident in this study.^38,39^ Increased phosphorylation of p38 and PARP by inhibition of *CPLX1* may reflect activated apoptotic response ^40^ Taken together, our result suggested that *CPLX1* promotes malignant phenotypes in gastric cancer cells, although the molecules, pathways, and their interactions with *CPLX1* should be investigated more.

Our epidemiological data suggested that *CPLX1* may be a biomarker predicting recurrence of stage II-III gastric cancer, especially peritoneal recurrence despite surgery followed by oral fluoropyrimidine. These high-risk patients may have a chance to undergo more intensive chemotherapy after surgery or even prior to surgery by measuring *CPLX1* expression in the primary tumor. Two adjuvant chemotherapy regimens of oral fluoropyrimidine (S-1) monotherapy and Capecitabine plus Oxaliplatin have been standard in East Asia so far.^41-43^ However, oral fluoropyrimidine monotherapy for stage III has been considered to be less effective than for stage II.^44^ Several clinical trials have been conducted to seek survival benefit from more intensive regimens than the monotherapy.^44,45^ We might be able to select a high-risk cohort who can benefit from those aggressive regimens by measuring *CPLX1* expression. We also might be able to develop a molecular-targeted drug for *CPLX1* combined with fluoropyrimidines.

The present study has certain limitations. First, the present data about *CPLX1* mRNA expression data was retrospectively collected from a single institution, and the cut-off value for high *CPLX1* expression was exploratorily defined. Second, mechanisms of how *CPLX1* promote malignant phenotypes of gastric cancer should be investigated more. In particular, the dynamic behavior of *CPLX1* and relation with cancer progression molecules in cancer cell should be further investigated. For example, visualizing that *CPLX1* protein (Complexin-1) and EMT-associated molecules are colocalized and demonstrating alteration of VC by si*CPLX1* in cancer cells would prove our postulation. Moreover, experiments with forced expression of *CPLX1* and in vivo experiments are preferable to be done to confirm that *CPLX1* promotes malignant phenotypes and plays a role in drug resistance.

In conclusion, increased expression of *CPLX1* in gastric tissues may be a prognostic biomarker predicting recurrence after curative resection of gastric cancer followed by adjuvant chemotherapy. In addition, *CPLX1* promotes malignant phenotypes and may be a therapeutic target to prevent the recurrence of gastric cancer.

## Data Availability

Data will be available as needed for reasonable purposes under the appropriate procedure.

## Abbreviations

EMT: epithelial-mesenchymal transition
mRNA: messenger RNA
OD: optical density
qRT-PCR: quantitative reverse-transcription polymerase chain reaction
RNAseq: RNA sequencing
siRNA: small interfering RNA
VC: vesicular cycle

## Acknowledgment

We thank Taiho Pharmaceutical Co. Ltd for technical support, advice on this project through discussion on our result.

---

**Supplemental Fig. S1.**
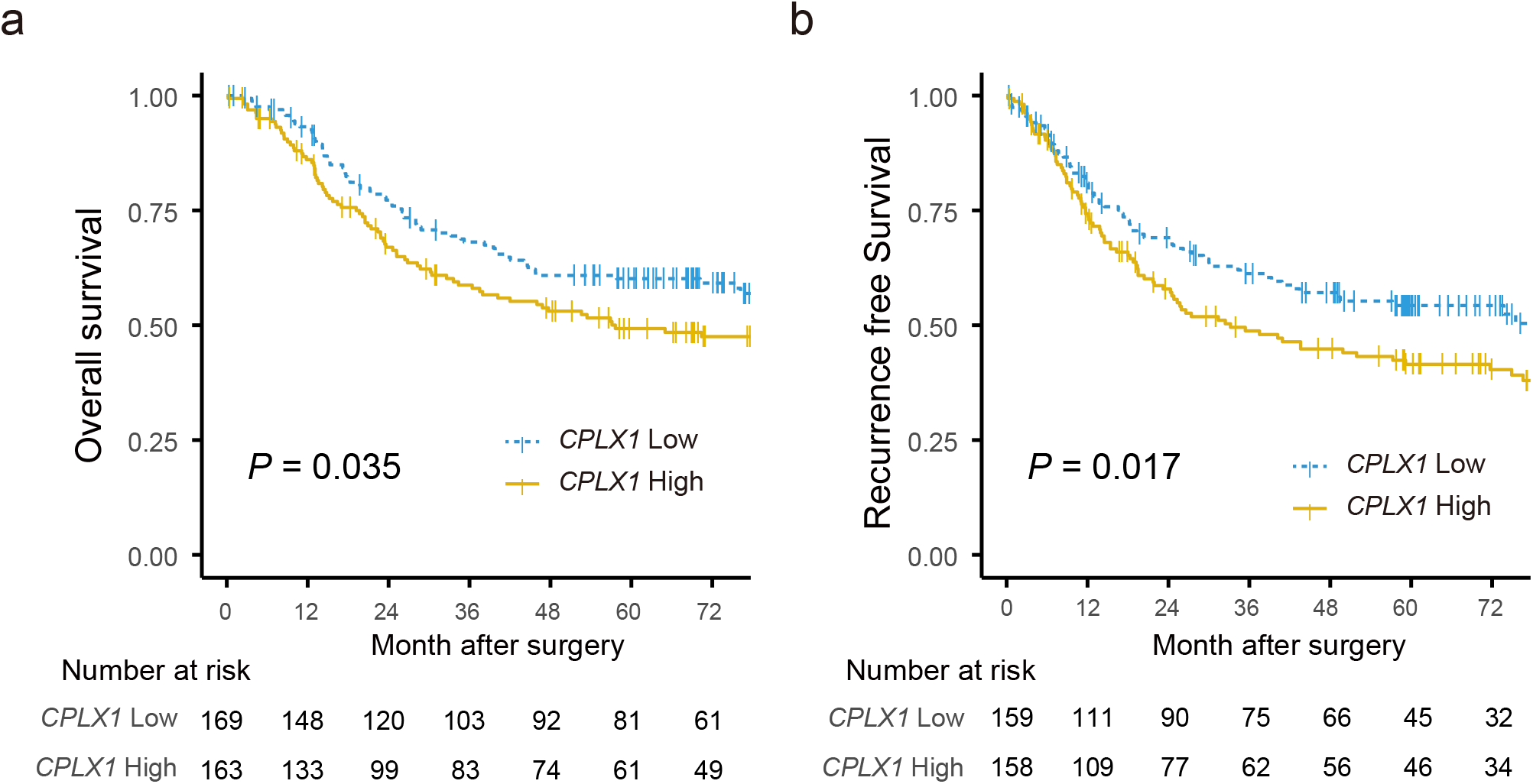
Kaplan-Meier plot of (a) overall survival, and (b) progression-free survival using external datasets grouped by the median value of CPLX1. Data were retrieved from the GSE62254, GSE15459, GSE22377, GSE62254, GSE29272, and GSE38749, of which UICC Stages were limited to II-III.

**Supplemental Fig. S2.**
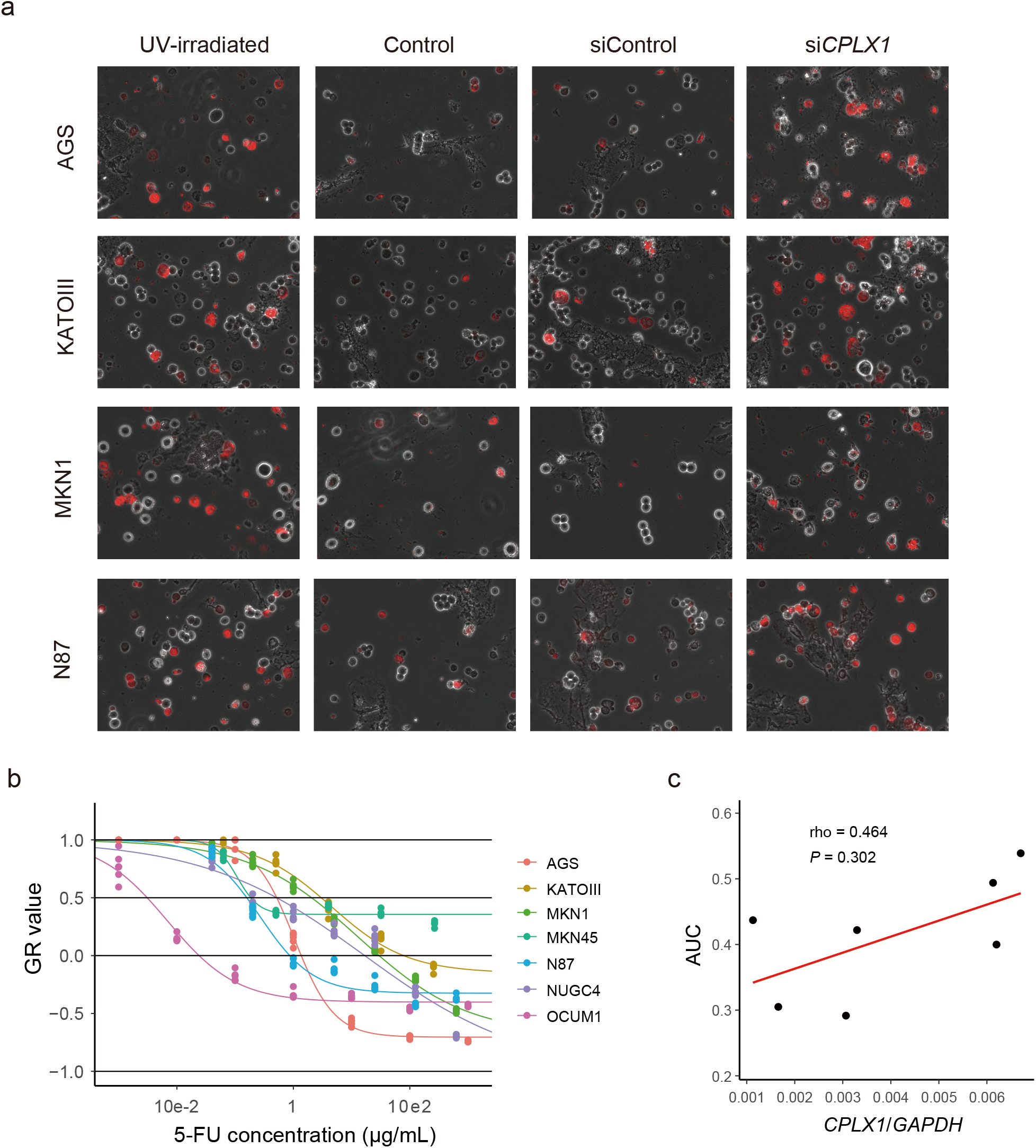
(a) Apoptotic cell detection by staining with annexin-V (See Fig. 2e). Phase-contrast and fluorescence images were merged to be presented. (b) Drug sensitivity tests to fluorouracil (5-FU) on seven gastric cancer (GC) cell lines. GR values indicate values calculated by normalized growth rate inhibition (GR) metrics. (c) Scatter plot between *CPLX1* mRNA expression (*CPLX1/ GAPDH*) and area under the dose-response curves (AUC) to fluorouracil (5-FU) of seven GC cell lines. A correlation test was done with the Spearman test.

**Supplementary Table S1.**
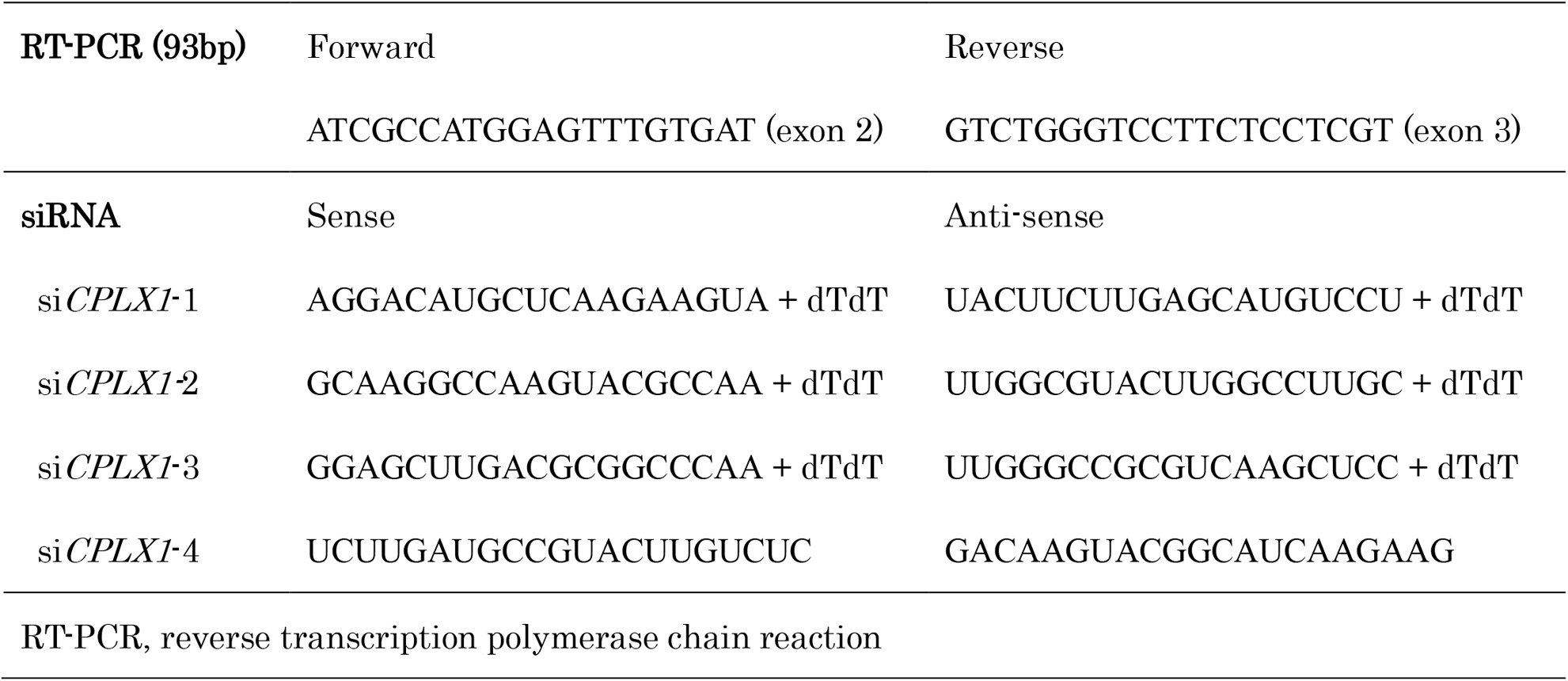
Primer sequences.

